# The association between type 2 diabetes disease trajectories and dementia incidence

**DOI:** 10.64898/2026.06.24.26356489

**Authors:** Scott C Zimmerman, Lucia Pacca, Whitney Wells, Sarah F Ackley, M Maria Glymour

**Affiliations:** Department of Epidemiology, Boston University School of Public Health, Boston, MA; Department of Epidemiology and Statistics, University of California San Francisco, San Francisco, CA; Department of Public Health, Washington University in St. Louis. St Louis, MO; Department of Epidemiology, Brown University, Providence, RI

## Abstract

**Introduction:** Type 2 diabetes (T2D) prevalence, severity, duration, and control are associated with dementia incidence, but prior literature is focused on specific pharmacologic, dietary, and exercise interventions in isolation while controlling for other co-occurring factors. Accounting for comprehensive life course experiences of the timing of diabetes onset, severity, treatment, and progression over a period of decades would provide a more comprehensive description of how life course diabetes progression and control is associated with dementia. Trajectories of diabetes diagnosis, pharmacological management, and disease progression are heterogeneous, and classifying these trajectories presents a significant methodological challenge.

**Methods:** Using deidentified survey and electronic health record data from Kaiser Permanente Northern California (KPNC) from the Research Program on Genes, Environment, and Health (RPGEH), we defined annual “states” for each eligible participant with T2D diagnosed between ages 50 and 70 based on KPNC, diabetes diagnosis, glycated hemoglobin, antidiabetes prescription count, and kidney dysfunction. We then employed sequence and cluster analyses to group participants into clusters with similar trajectories of these states. Finally, we estimated hazard ratios for incidence of Alzheimer’s disease and Alzheimer’s disease related dementias (AD/ADRD) for each of these clusters as well as individuals with type 1 or other diabetes types, relative to participants without diabetes at age 70, using covariate-adjusted Cox proportional hazards models.

**Results:** Using the 18,688 participants with T2D included in the diabetes trajectory assessment, sequence and cluster analysis identified 9 clusters of T2D treatment and control histories between ages 50 and 70. Clusters differed markedly in timing of onset of T2D, glucose control, antidiabetes drug use and kidney function. Associations of these clusters with incident AD/ADRD after age 70 was heterogeneous and patterned by diabetes control and treatment history, particularly by diabetes duration and treatment regime.

**Conclusions:** In conclusion, in this real-world data context, we find increased diabetes severity, increased medication use, and faster progression to kidney disease is associated with increased risk of dementia. We find some patterns of diabetes severity and control are associated with greater dementia risk. This information may be useful in the context of targeted screening and allocation of preventative services for ADRD.

## Introduction

Type 2 diabetes (T2D) prevalence, severity, duration, and control are associated with dementia incidence (Lee 2019, Reinke 2022, Yaffe 2012, Campbell 2018). Significant increases in prevalence of T2D in older adults (Selvin 2014), changes in life expectancy, and robust associations between glucose control and dementia suggest that optimizing glucose control and drug treatment regimens represents a significant opportunity to reduce dementia incidence (Crane 2013, González-Reyes 2016, ADA 2023, CDC 2023). Prior literature has focused on the shorter-term effects of specific pharmacologic, dietary, and exercise interventions in isolation while controlling for other co-occurring factors (Pelle 2023, Sun 2023, Tang 2023, Wang 2022, Yoo 2022, Chou 2017). Accounting for comprehensive life course experiences of the timing of diabetes onset, severity, treatment, and progression over a period of decades would provide a more comprehensive description of how life course diabetes progression and control is associated with dementia.

Elevated blood glucose, as measured by glycated hemoglobin (HbA1c), is a marker of diabetes severity (Sherwani 2016). High blood glucose may increase dementia risk via the direct effects of blood glucose on the formation of advanced glycation end products, inflammation, hyperlipidemia, and microvascular disease, or through the development of diabetes complications that are associated with increased dementia risk (Moran 2023).

Control of blood glucose is a major goal of diabetes treatment, and may be obtained through lifestyle changes or pharmacological treatment. Prior work examining HbA1c trajectories found that poor HbA1c control following diagnosis can increase the risk of diabetes complications associated with increased dementia risk (Laiteerapong 2017). Lifestyle changes such as smoking cessation, diet and exercise can help control HbA1c and are associated with deceased dementia risk (Barnes 2015, Demurtas 2020, Gungabissoon 2022). However, control of blood glucose using antidiabetic agents may influence dementia risk in individuals with diabetes via processes beyond their glucose lowering effects (Campbell 2018). For example, metformin (N,N-dimethylbiguanide) is very effective at lowering blood glucose and decreasing insulin resistance (Landin 1991, Dean 2016, Horakova 2019, Zhou 2001), and is associated with decreased dementia incidence (Sun 2023, Zimmerman 2023). On the other hand, specific antidiabetic agents may be harmful with respect to cognitive aging: insulin may be uniquely harmful with respect to dementia outcomes since it is associated with cardiovascular disease (Pyörälä 2000, Rensing 2011), and thus may contribute to dementia risk via its negative vascular effects (Muniyappa 2007).

Complications may occur when diabetes control is not obtained. One serious complication, kidney dysfunction (Bloomgarden 2005), is particularly strongly associated with dementia incidence (Ghoshal 2020), and harmful interactions between kidney dysfunction and diabetes drugs are known to occur (Sommer 2020).

Trajectories of diabetes diagnosis, pharmacological management, disease progression, and related complications are complex and heterogeneous, and classifying these trajectories presents a significant methodological challenge. Sequence analysis (Abbott 1995) is a method to compare and categorize trajectories of lifecourse exposures such as educational attainment or employment history in order to evaluate the associations between these exposure trajectories and disease outcomes, including dementia (Vable 2020 & 2021, Duarte 2022). Sequence analysis has also been used to characterize trajectories of multimorbidity–including diabetes–and examine their association with hospitalizations and deaths (Cezard 2022). Among the main advantages of sequence analysis are its applicability to complex longitudinal data and its ability to account for timing, duration, and order of events.The characterization of diabetes trajectory patterns associated with dementia risk would enable increased screening and early intervention among higher-risk patients. Prescribing patterns identified from these exposure trajectory clusters may also allow for the identification of potentially modifiable factors that could be used to optimize diabetes treatment regimens for the prevention of dementia.

In this analysis we used sequence analysis to classify diabetes trajectories from age 50-69, and then evaluated the association between these trajectories and subsequent dementia risk.

## Methods

### Overview

In a large cohort of diabetes- and dementia-free individuals at age 50, we used sequence analysis to classify trajectories from age 50 to 69 based on annual states defined from: count of antidiabetic drug prescriptions, HbA1c control, and kidney function. We then use time-to-event analysis to determine the association of these trajectories with dementia incidence in participants who were dementia-free at the start of the outcome assessment period (age 70).

### Population and Setting

Kaiser Permanente Northern California (KPNC) is an integrated healthcare delivery system serving over 3.2 million adult members from a catchment area of 22 counties in northern and central California. Compared to insured adults aged 26-84 in the KPNC catchment area, KP members were similar with regards to race/ethnicity, educational attainment, and health and well-being indicators but were less likely to be covered by the California Medical Assistance Program (Medi-Cal), receive food stamps, or be in a lower income (<$35,000) household (Gordon 2020).

### Sample Data

This study used deidentified survey and electronic health record (EHR) data from Jan. 1 1996 to Dec. 31 2019 for KPNC members born before January 1, 1955 (and thus at least 65 years old by the end of the study period) who completed one of two harmonized health surveys: (1) the California Men’s Health Study (CMHS; Enger 2006), offered in 2002-2003 to men who were 45-69 years old in 2000 and who had been KPNC members for at least 1 year, or (2) the Research Program on Genes, Environment, and Health (RPGEH) survey, offered in 2007-2009 to people 18+ years old who had been KPNC members for at least 2 years. The EHR data consisted of detailed data on KPNC membership, diagnoses, laboratory tests, prescription dispensation records, diabetes and renal registry data, and death records. The survey data included self-reported health, sociodemographic and behavioral variables (See Methods Supplement).

Conceptually, we divided the life course into an exposure trajectory period (age 50-69) and an outcome assessment period (age 70-90). We restricted the sample to individuals who were alive and KPNC members at the end of the exposure period (see figure 1) and who were KPNC members for at least 10 years during this period to allow for adequate definition of exposure trajectories. To ensure the most complete characterization of patient exposure trajectories we retained patients who were diagnosed with dementia after the start of the exposure period in the exposure assessment stage, but we then excluded those individuals from the outcome assessment stage.

**Figure 1:**
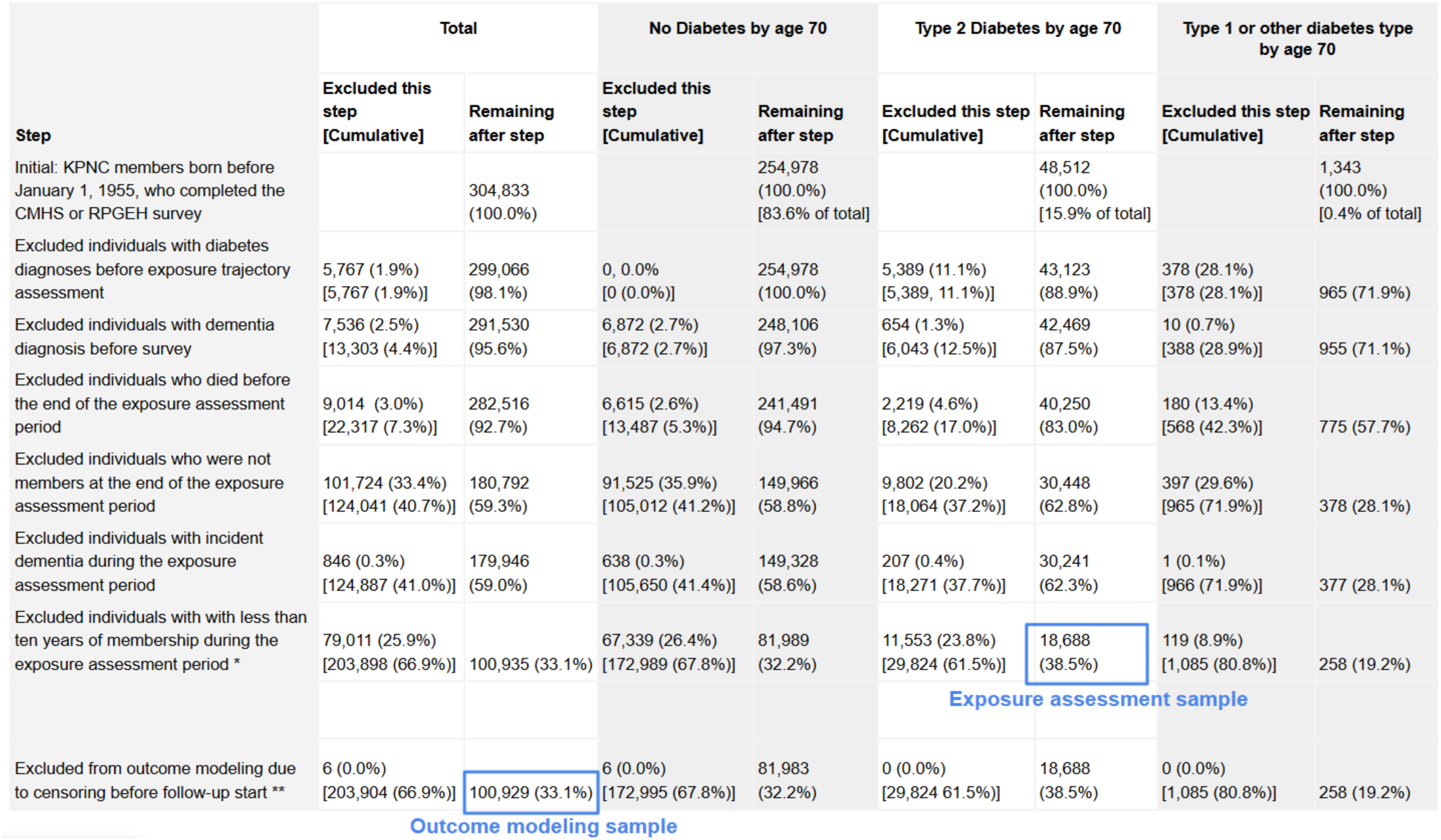
Sample construction.

### Measures

#### Yearly exposure states

We defined annual exposure states from age 50 to 69 based on KPNC membership, diabetes diagnosis, HbA1c, antidiabetes prescription records, and kidney dysfunction markers. KPNC membership was determined from administrative records based on whether the participant was enrolled at any point in the year. An individual’s first recorded diabetes diagnosis was determined based on the KPNC diabetes registry, and participants were classified as having a history of diabetes in a given year if their first diagnosis occurred before the end of that year. Antidiabetes prescription count was defined from five classes of oral hypoglycemic agents (OHAs: biguanides, sulfonylureas, alpha-glucosidase inhibitors, thiazolidinediones and meglitinides) and insulin of any type (Appendix Table 2). For each prescription class, we created indicator variables for whether the participant had a dispensation record for that drug class in that year. We added the indicators for the six drug classes to define a count of the total number of classes. We calculated the maximum recorded HbA1c in each year and then created classifications of HbA1c based on ranges (Controlled <=7%, High: 7%+); ADA 2022; Chobanian 2003), and filled in missing values by carrying forward previous values. We defined an indicator of kidney dysfunction based on any of the following: low eGFR (<= 60 mL/min/1.73m^2^), high creatinine (≥1.4 mg/dL for women and 1.5 mg/dL for men), diagnosis of kidney disease, or recorded dialysis. Missing lab values were coded as no kidney dysfunction.

#### Outcome and censoring dementias

Age of first diagnosis of Alzheimer’s disease and Alzheimer’s disease related dementias (AD/ADRD) was determined from the EHR based on ICD-9 and ICD-10 diagnosis codes for Alzheimer’s Disease, vascular dementia, and nonspecific dementia. See Appendix Table 1 for ICD codes.

#### Covariates in time-to-event analysis

Additional baseline covariates were included in time-to-event analysis models. Time-invariant sociodemographic data included sex (male, female), race/ethnicity (Asian, Black, Hispanic, White, missing/other), foreign birth (yes, no, unknown), each parent’s foreign birth (yes, no, unknown), survey language (English, Spanish, Chinese), education (less than high school, high school graduate, college graduate or greater, and other), marital status (married or living as married: yes/no), and category of annual income (<$20,000, $20,000-$39,999, $40,000-$59,999, $60,000-$99,999, or $100,000+).

#### Covariates in descriptive analysis and sensitivity analyses

In addition to variables above, we also provide comparisons of covariate distributions across clusters at each decade from 50-90 years of age. Since there was a large amount of missingness in these covariates prior to age 50, we did not include these as potential confounders in the time-to-event models. The covariates included: body mass index; blood pressure; use of nondiabetes medications (antilipemics, hypertension drugs, cardiovascular disease (CVD) drugs); indicators of diagnosis and continuous covariates for time since diagnosis of comorbidities (hypertension, cancer); count of CVD event categories (LIST). ICD codes for EHR variables are presented in Appendix Table 1, and pharmaceutical codes are presented in Appendix Table 2.

In order to protect against birth date identification, each participant was assigned an “administrative censoring” date randomly chosen to be between January 30th, 2020 and March 31st, 2020. Ages were topcoded at age 90 during the data deidentification process.

### Statistical Analysis

#### Overview

We used sequence analysis and cluster analysis to define clusters of similar participants among participants who developed diabetes before age 70. We then pooled these participants with those who did not develop diabetes by age 70 (assigned to their own cluster), and performed time-to-event analysis for the outcome assessment period to compare hazards of dementia for each cluster using Cox proportional hazards modeling (Appendix Figure 2a).

**Figure 2:**
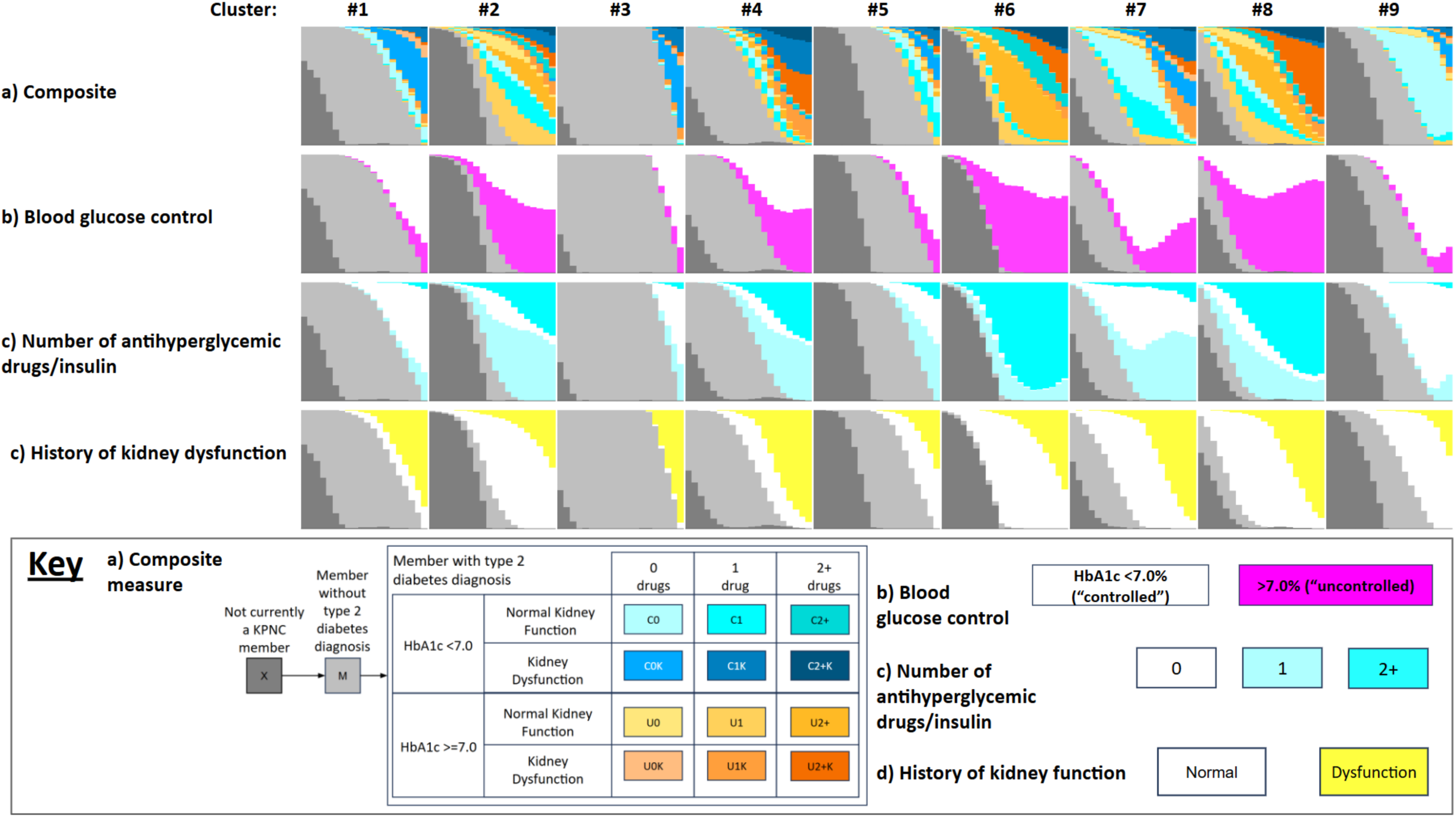
Patterning of blood glucose control, number of antidiabetes drugs & insulins, and history of kidney dysfunction from age 50-70 across clusters identified by sequence and cluster analysis. Density plots for 9 clusters of diabetes history from age 50-70. The x axis of each sub-plot spans ages 50-70, and the y axis denotes the proportion of the cluster at each age that falls into each category. Row (a) describes the composite measure with 14 states defined by KPNC membership, history of T2D diagnosis, and, for those with a history of T2D diagnosis 12 states defined by combinations of: blood glucose control, number of antihyperglycemic drugs & insulin, and history of kidney dysfunction. Rows (b)-(d) illustrate the densities of each of these three components of the composite measure for people with a history of a T2D diagnosis. KPNC: Kaiser Permanente of Northern California; T2D: Type 2 diabetes; HbA1c: Hemoglobin A1c.

#### Sequence and Cluster Analysis

For participants who developed diabetes before age 70, we conducted sequence analysis on the exposure trajectory period (ages 50-69) to classify participants with similar exposure state trajectories into clusters.^28^ Inclusion of participants who did not develop diabetes before age 70 in the cluster analysis was computationally infeasible, and since the only variation within this group would be due to membership, we defined this group as a cluster of interest a priori and excluded it from the sequence and cluster analysis.

Defining the trajectories consisted of three steps. First, individual trajectories (also called sequences) were created by assigning each year from age 50 to age 69 to a state based on a combination of membership (member or not a member) and diabetes diagnosis history (not yet diagnosed or diagnosed). For those with a history of diabetes diagnosis 12 states were constructed based on the combinations of diabetes control (HbA1c < 7% vs 7%+), count of classes of OHA and insulin use (none, 1 class, 2 or more classes), and kidney dysfunction (yes/no). Thus our categorization produced a total of 14 possible states (Figure 2). Appendix Figure 2b illustrates several hypothetical exposure trajectories that might be observed during the exposure trajectory.

Second, we used the optimal matching algorithm, which prioritizes duration and ordering of states in the sequences, to define a matrix of dissimilarity costs between each individual trajectory and all the other trajectories in our data (Abbott 2000). The dissimilarity costs between each pair of trajectories were obtained by summing insertion/deletion (“indel”) and substitution costs corresponding to replacing states that differed between the two trajectories at each time point. Substitution costs were based on observed transition frequencies between states in our data such that fewer transitions corresponded to higher costs. Indel costs were set to be 0.5 times the highest substitution cost.

Finally, we used agglomerative, hierarchical cluster analysis with Ward linkage (Ward 1963) to group similar trajectories based on the dissimilarity costs matrix. The final number of clusters was chosen based on the Duda-Hart cluster stopping rule (Duda 1973) as well as cluster interpretability and minimum cluster sample size. In sensitivity analysis we repeated these steps using the Dynamic Hamming dissimilarity measure (Lesnard 2010), which prioritizes timing instead of duration and ordering of states. Participants were coded with a “non-member” state for time points at which they were not members.

We summarized sequence analysis results using index plots (individual-level trajectories) and chronograms (summary of states distribution over time) for each of the identified clusters. In addition, we compared descriptive plots and statistics of each cluster of diabetes trajectories, and fit statistics for each choice of number of clusters.

#### Time-to-event analysis

We used Cox proportional hazards models to estimate the association between exposure trajectory cluster and ADRD incidence. If participants were not diagnosed with diabetes between age 50 and 69 (and thus not included in the exposure sequence analysis), they were assigned to the reference category of “no diabetes”. Additionally, if participants would have been included in the exposure assessment except for the fact that they were diagnosed with Type 1 diabetes or diabetes of unknown type they instead were included in an additional “Type 1/other diabetes” exposure category.

Participants were followed from “baseline”, defined as age 70 or age of survey (whichever occurred later), to avoid inclusion of immortal person-time for those who completed surveys after age 70 (Platt 2019). For participants with multiple membership periods, membership periods were collapsed if there was less than a year gap between them. The end of the collapsed membership period that spanned the age baseline was used for censoring. End of follow-up was defined as the first of: ADRD diagnosis, death, reaching age 90, end of the KPNC membership period the participant was in at baseline, or the end of the study period (Dec. 31, 2019). (Figure 2b)

Participants were excluded from the outcome time-to-event analysis if they had an AD/ADRD diagnosis, death, turned age 90, end of membership, or end of the study period before the start of the outcome assessment follow-up period. The exposure trajectory cluster was treated as a categorical variable with diabetes-free at baseline as the reference category.

We estimated Cox models adjusting for three sets of covariates. Model 1 adjusted for basic demographics (sex, race/ethnicity). Model 2 additionally adjusted for time-invariant survey measures plausibly reflecting covariates prior to the beginning of the exposure period (foreign birth, parents’ foreign birth, education, survey language). Model 3 additionally included marital status at time of survey (See Table 2).

Analyses were conducted using R version 4.3.1. Sequence analysis and cluster analysis were performed using the TraMineR, fastcluster and WeightedCluster libraries in R. Human subjects approval was granted by the University of California San Francisco, and the Kaiser Permanente Northern California, Mid-Atlantic States Institutional Review Boards. We followed the Strengthening the Reporting of Observational Studies in Epidemiology (STROBE) reporting guidelines (Elm 2007).

## Results

Of an initial sample of 304,833 participants, 48,512 were diagnosed with T2D by age 70, and 1,343 were diagnosed with T1D or other diabetes, and 254,978 had no diabetes diagnosis by age 70. After exclusions, the final analytic sample for exposure assessment consisted of 19,699 participants with T2D by age 70, and the final outcome modeling sample included 100,929 participants (81,983 [81.2%] without diabetes; 18,688 [18.5%] with T2D; and 258 [0.3%] with T1D or other diabetes by age 70; Figure 1).

### Exposure assessment: Sequence and cluster analysis among participants with a T2D by age 70

Using the 18,688 participants included in the exposure assessment, sequence and cluster analysis identified 9 clusters of T2D treatment and control histories between ages 50 and 70. The most common exposure sequence among those with diabetes before age 70 was characterized by high HbA1c despite use of 2+ antidiabetes medications, and moderate history of kidney dysfunction (Cluster #8). Clusters #6 and #8 were defined by early onset of T2D and extended periods of high HbA1c and high use of antidiabetes medications, and differed in terms of time with kidney dysfunction (#2: low, #8: moderate). Clusters #2 and #4 also had high proportions of time with uncontrolled diabetes, but lower use of antidiabetes drugs. Cluster #2 had later T2D onset and lower levels of kidney dysfunction than #4. Cluster #7 was similar in T2D onset timing to cluster #2 but with lower levels of uncontrolled HbA1c and lower use of antidiabetes drugs, but higher levels of kidney dysfunction. Clusters #1 and #3 had later average onset of T2D, with #1 slightly earlier than #3. Both had low levels of uncontrolled glucose and antidiabetes drug use but high levels of kidney dysfunction. Cluster #9 had low levels of uncontrolled HbA1c with low antidiabetes drug use and low levels of kidney dysfunction. Cluster #8 had the highest proportion of years with simultaneous uncontrolled glucose, high antidiabetes drug use, and kidney dysfunction history, followed by clusters #4 and #6 (Figure 2).

### Outcome assessment: Associations between T2D histories by age 70 and subsequent dementia incidence

3.4% of participants without diabetes diagnosis before outcome assessment baseline (the latter of age 70 and age at survey) received a dementia diagnosis during subsequent follow-up, while the proportion was higher for those with T2D (6.2%) and T1/other diabetes (5%). 3.0% of participants without diabetes at baseline developed diabetes during follow-up. Mortality was lower for participants without diabetes (9.5%, mean age 73.8 years) compared to those with T2D (15.8%, mean age 74.5 years) or T1/other diabetes (19.0%, mean age 73.9 years) at baseline. Those with T2D were less likely (45.7%) while those with T1/other diabetes were more likely (54.7%) to be female than those without diabetes at baseline (47.7%). Across the identified clusters of T2D histories from exposure assessment, sociodemographic indicators varied substantially (Table 1).

**Table 1.**
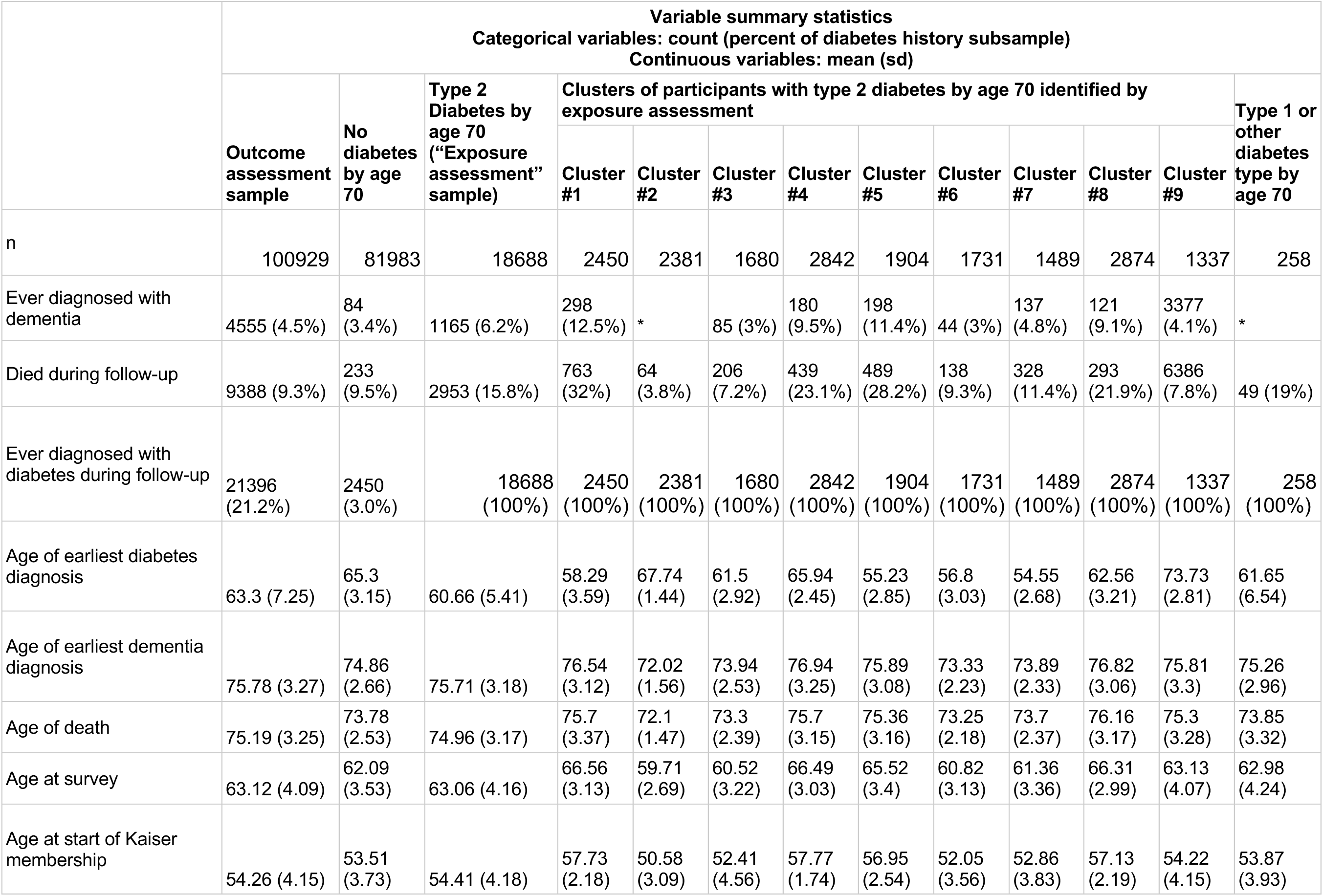
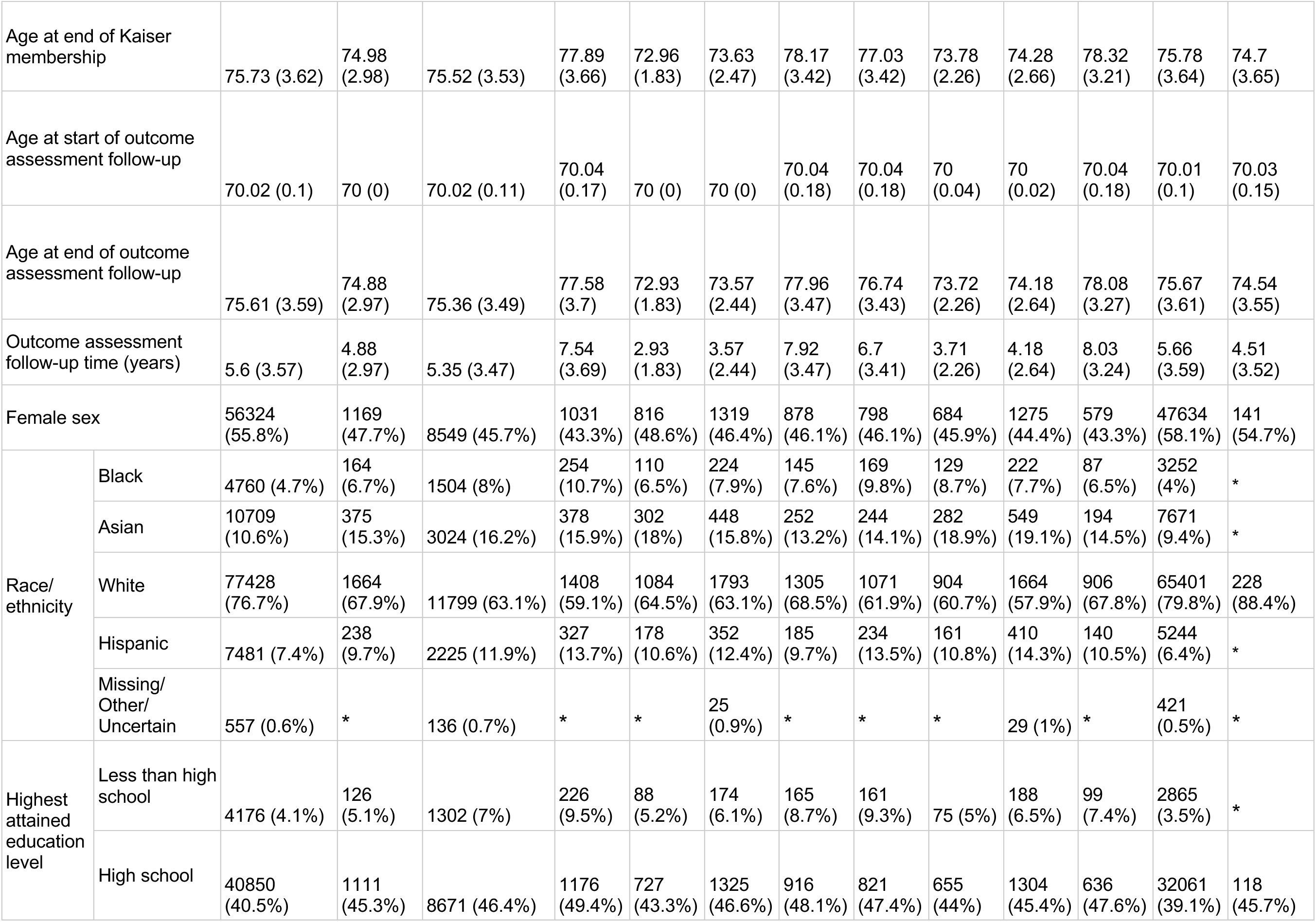
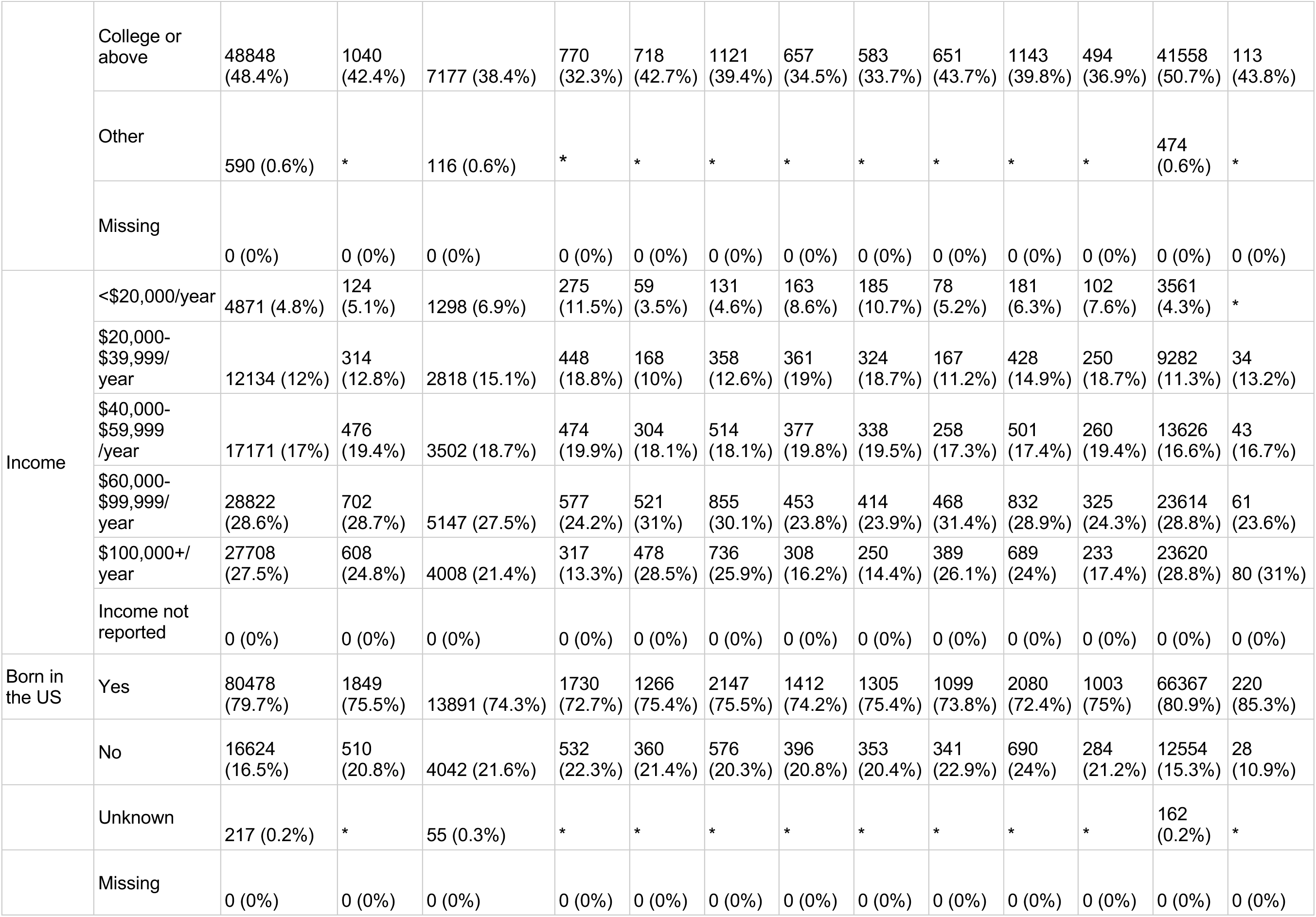
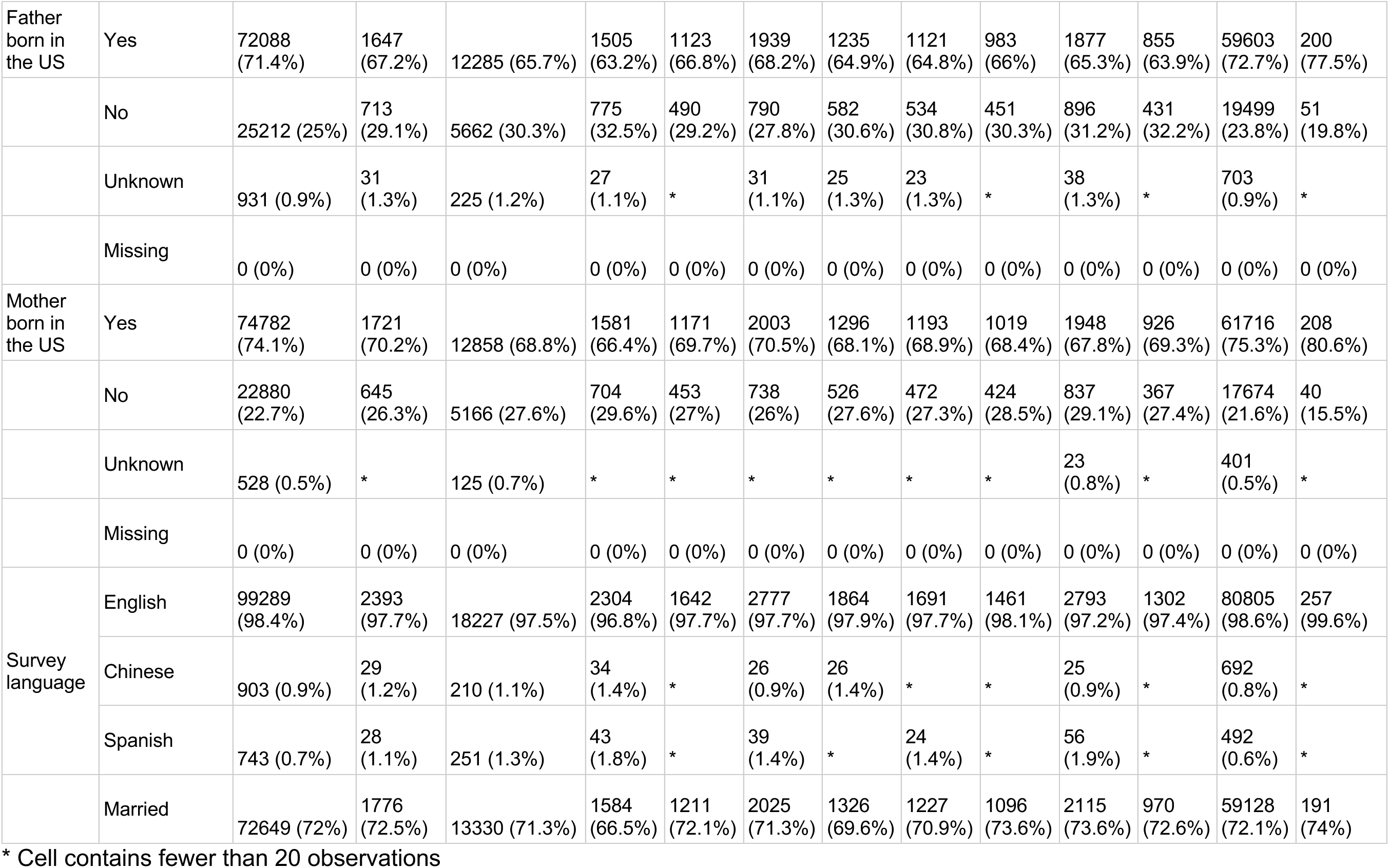
Sample characteristics.

Table 2 and Figure 3 show the hazard ratios for dementia by exposure trajectory with individuals not diagnosed with diabetes before baseline as the reference group. Using the 81,983 participants included in the outcome assessment, hazard ratios for participants with a history of T2D by age 70 clustered by history of blood glucose control, antidiabetes drug use and kidney dysfunction ranged from 0.77 (95%CI: 0.46-1.28; #3) to 2.29 (95%CI: 1.97-2.66; #6) relative to the group without diabetes at baseline when adjusting for covariates plausibly occurring prior to the exposure period. Increased covariate control had minimal effect on the estimated hazard ratios. The lowest hazard was for cluster #3, which was primarily characterized by the latest onset of T2D. The highest hazard ratio was for cluster #6, which was characterized by relatively early onset of T2D and long periods of uncontrolled blood glucose with high antidiabetic drug use. Clusters #2 – characterized by onset of T2D around age 60 and high portions of time with uncontrolled blood glucose with moderate-to-high use of antidiabetic drugs and low portions of time with kidney dysfunction – and #8 – characterized by early onset of T2D and extended periods of high HbA1c and high use of antidiabetes medications and moderate time with kidney dysfunction – also had high hazard ratios relative to those without diabetes (#2 HR=1.92 (95%CI: 1.69-2.18); #8 HR=1.91 (95%CI: 1.58-2.31)). The hazard ratio for T1/other diabetes was elevated but confidence intervals crossed the null due to the small sample size (95%CI: 0.46-1.28).

**Figure 3.**
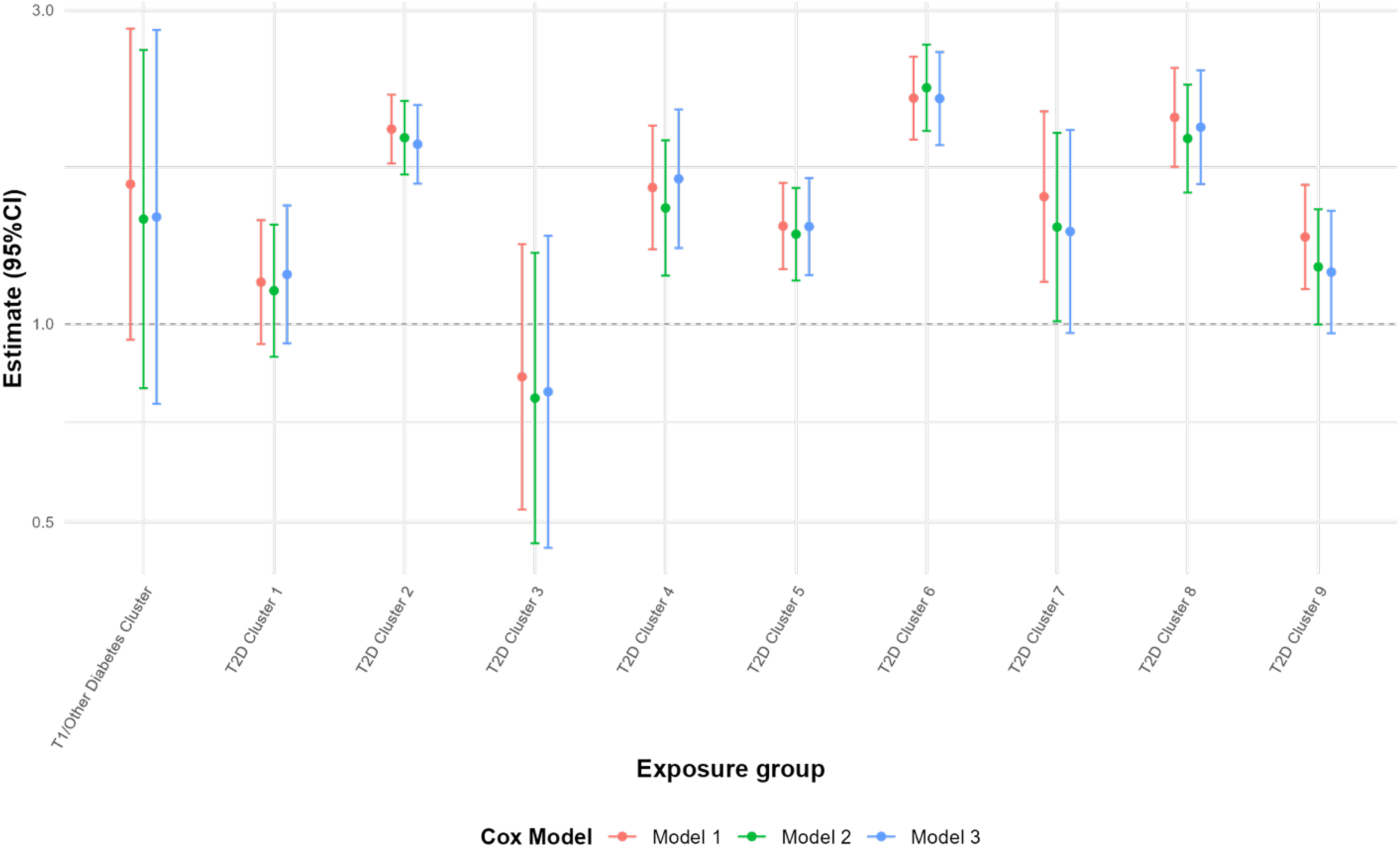
Hazard ratios for Alzheimer’s disease & Alzheimer’s disease across diabetes histories at age 70 compared to those without diabetes at age 70. The exposure was treated as a 11 categorical variable (diabetes history by age 70: none (reference), T2D clusters 1-9, and T1 or other diabetes type) with no diabetes by age 70 as the reference category. Clusters 1-9 included participants with T2D by age 70 analyzed using sequence and cluster analyses to group participants based on patterns of diabetes status, KPNC membership, blood glucose control, kidney function, and diabetes drug use in each year from ages 50-70 (see Figure 2). We estimated Cox models adjusting for three sets of covariates. Model 1 adjusted for basic demographics (sex, race/ethnicity). Model 2 additionally adjusted for time-invariant survey measures plausibly reflecting covariates prior to the beginning of the exposure period (foreign birth, parents’ foreign birth, education, survey language). Model 3 additionally included marital status at time of survey. KPNC: Kaiser Permanente of Northern California; T2D: Type 2 diabetes.

**Table 2.**
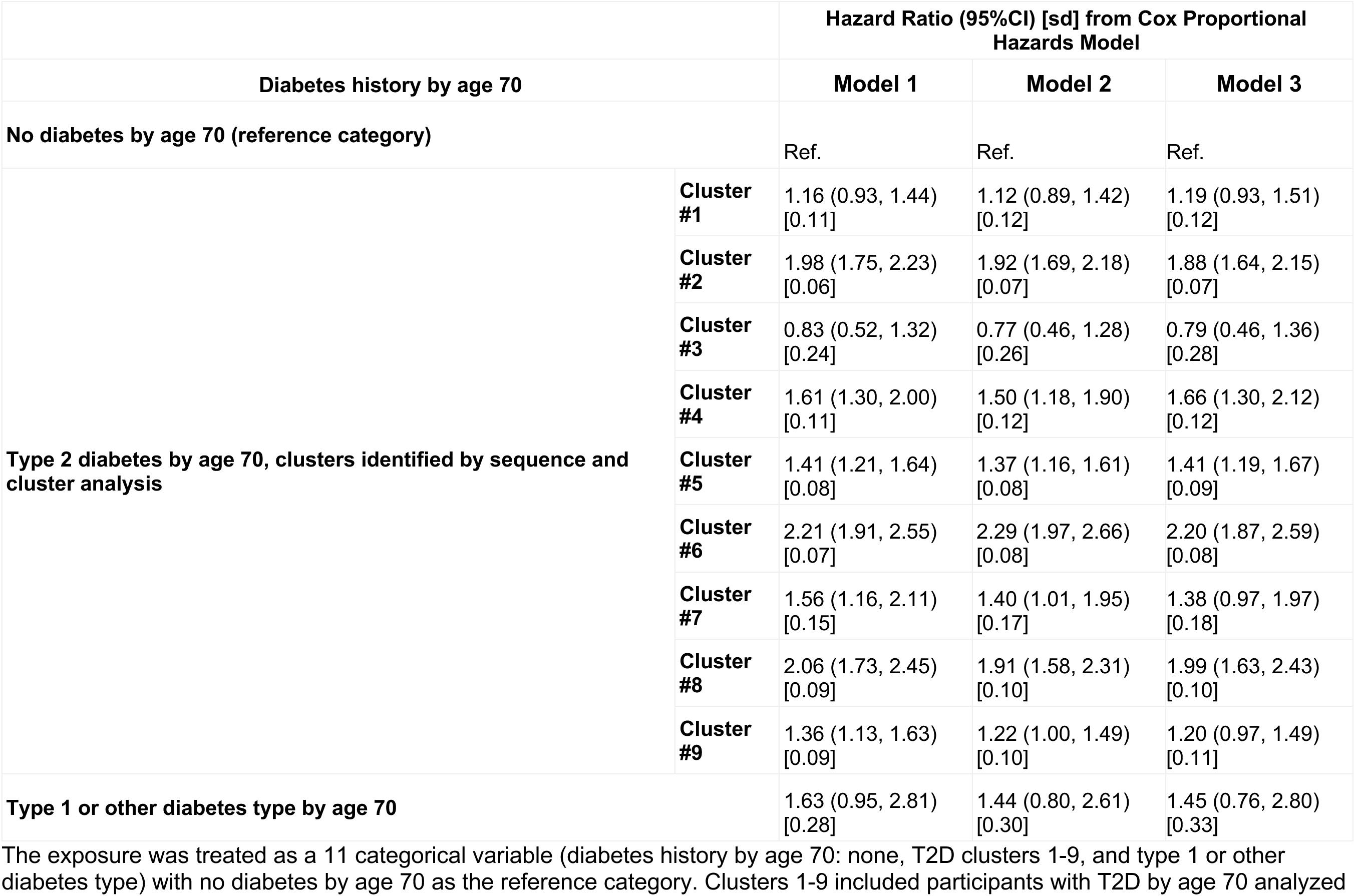
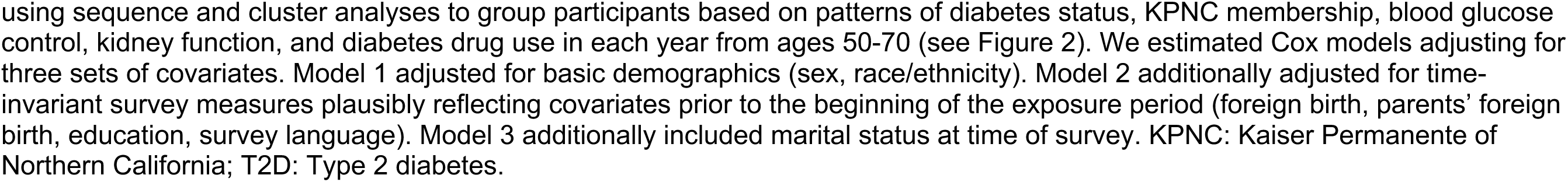
Hazard ratios for Alzheimer’s disease & Alzheimer’s disease across diabetes histories at age 70 compared to those without diabetes at age 70.

## Discussion

In a cohort of 18,688 eligible members of KPNC diagnosed with T2D by age 70, sequence and cluster analysis grouped together participants with similar histories of diabetes treatment and control between ages 50 and 70. The resulting clusters illuminate how timing of diabetes onset, treatment and control are associated with incidence of AD/ADRD after age 70.

Early onset of T2D was particularly strongly associated with AD/ADRD incidence, with later onset T2D (Clusters #1 and #3) demonstrating similar hazards of dementia compared to those without diabetes at age 70. Likewise, diabetes control plays an important role. For example, T2D with onset around age 60 was less associated with AD/ADRD incidence after age 70 among those with better controlled diabetes with few drugs (cluster #9) compared to those with less control despite higher use of antidiabetic drugs (#4). The cluster with the highest proportion of time with uncontrolled diabetes (#6) also had a very short time between initiation of KPNC membership and T2D diagnosis, suggesting that T2D was present prior to initiation of KPNC membership.

These results add to previous research (Albai 2019) detailing heterogeneity in dementia risk for individuals with type 2 diabetes depending on diabetes control and treatment history, particularly by diabetes duration and treatment regime.

This analysis has the following strengths. The large and diverse sample has extended follow-up, allowing ample time for classification of exposures and identification of AD/ADRD cases. Furthermore, the EHR record includes detailed lab and diagnostic information that richly describes each participant’s history of diabetes treatment and control prior to age 70.

This analysis has the following limitations. First, since this analysis is descriptive using data from a real-world context, causal effects should not be attributed to the clusters identified. Limited available data about participant health and behaviors prior to age 50 restricted our ability to control for confounding. Each cluster represents a dynamic interplay between diabetes severity, treatment, complications and side effects over time. Diabetes is a heterogeneous disease (Pearson 2019), where disease progresses more quickly in some individuals, requiring additional medications earlier to effectively manage blood glucose levels. Additionally, responses to antidiabetic drugs differ across individuals. As such, differences between clusters may represent both differential benefits of treatment regimens and confounding by indication in which participants take more medication as a result of more severe or treatment-resistant diabetes. Second, because not all participants joined KPNC at the same time, our sequences include missing states. Some of the participants only joined the Kaiser health system after age 50, therefore we do not have information on their diabetes diagnosis and treatment for the first part of their trajectories. Handling missing values is a challenge in sequence analysis, since it may lead to biases in the final cluster solution due to the creation of “artificial” clusters (Lazar 2017). Moreover, excluding participants who ended KPNC membership may lead to selection bias, since participants who changed or canceled their health plan may have different health compared to those who stayed in the Kaiser system. Third, there is the possibility for measurement error for diagnoses in EHR data which are only recorded as clinically indicated. Fourth, Our analysis used age 50-70, this was chosen a priori based on data availability and the early edge of the distribution of ADRD onset, and may not be the most important time period, nor does it allow for classification of dementia risk. Further work should extend these findings to allow for age-specific predictions of dementia incidence of clinical utility. Finally, time-to-event analyses have the potential for selection bias since only individuals surviving and dementia free at the start of the outcome assessment period were included. Patterns in exposure trajectories may partially reflect preclinical and undiagnosed ADRD, which may begin decades before ADRD symptom onset and diagnosis.

In conclusion, in this real-world data context, we find increased diabetes severity, increased medication use, and faster progression to kidney disease is associated with increased risk of dementia. We find some patterns of diabetes severity and control are associated with greater dementia risk. This information may be useful in the context of targeted screening and allocation of preventative services for ADRD.

## Data Availability

The RPGEH survey data and GERA genotype data are available to qualified researchers by application to the Kaiser Permanente Research Bank, contingent on project approval by the Kaiser Permanente Research Bank Access Review Committee, institutional review board approval, and execution of a Materials and Data Transfer Agreement.

## Appendix

**Appendix Figure 2a:**
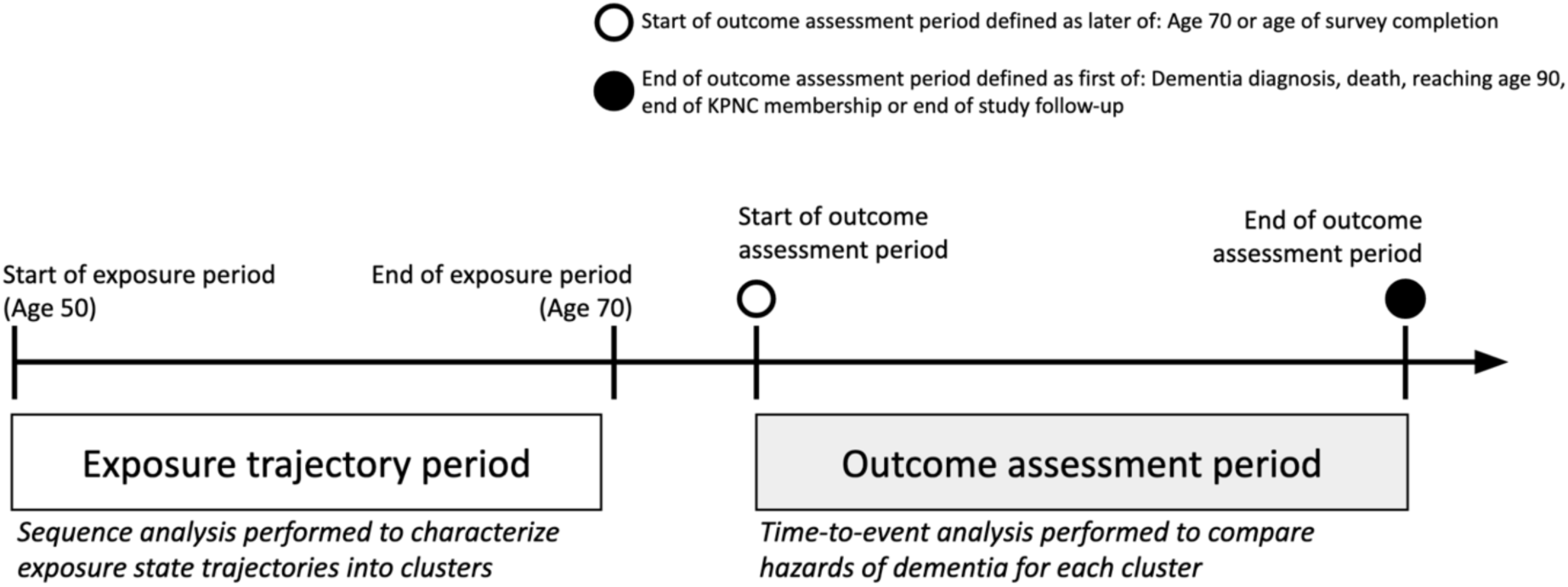
Summary of study periods. Illustration of the analytical approach, describing the criteria that determines the start and end point of each analytical period, and the statistical approach used in each analytical period. Abbreviations: Kaiser Permanente of Northern California (KPNC).

**Appendix Figure 2b:**
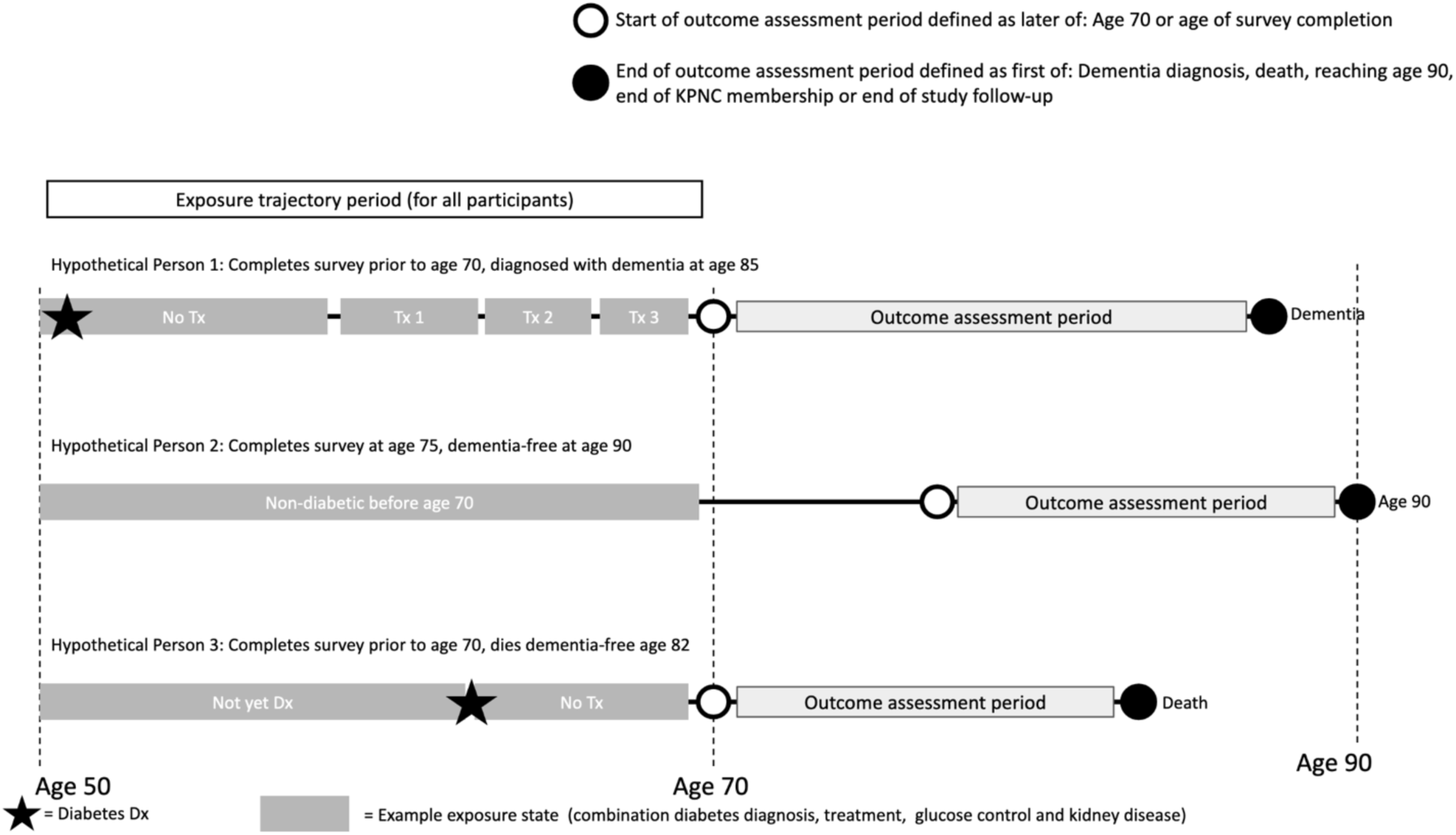
Example exposure trajectories and outcome assessment periods. Illustration of exposure trajectory periods and outcome assessment periods for hypothetical individuals, depicting possible sequence trajectories and possible start and end points for the outcome assessment periods depending on individual characteristics. Abbreviations: Kaiser Permanente of Northern California (KPNC), Treatment (Tx), Diagnosed (Dx).

**Appendix Table 1:** ICD codes.

| Diagnosis | ICD-9 codes | ICD-10 codes |
| --- | --- | --- |
| **Outcomes** |  |  |
| Alzheimer's Disease | 331.0 (Alzheimer's disease) | G30.x (Alzheimer's disease) |
| Vascular Dementia | 290.4x (Vascular dementia) | F01.5x (Vascular dementia) |
| Nonspecific Dementia | 290.0, 290.1x, 290.2x, 290.3, 294.2x, 294.8 (Nonspecific dementia) | F03.9x (Nonspecific dementia) |

**Appendix Table 2:** Prescription codes.

| Pharmacy drug class (AHFS) | Generic names |
| --- | --- |
| Oral Hypoglycemic Agents |  |
| 68:20.20 Sulfonylureas | Chlorpropamide |
|  | Glimepiride |
|  | Glipizide |
|  | Glyburide |
|  | Tolazamide |
|  | Tolbutamide |
| 68:20.04 Biguanides | Metformin Hydrochloride |
| 68:20.28 Thiazolidinediones | Pioglitazone Hydrochloride |
|  | Rosiglitazone Maleate |
| 68:20.16 Meglitinides | Nateglinide |
|  | Repaglinide |
| 68:20.02 α-Glucosidase Inhibitors | Acarbose |
|  | Miglitol |
| Insulins |  |
| 68:20.08.04 Rapid-acting Insulins | Insulin Aspart |
|  | Insulin Glulisine |
|  | Insulin Lispro |
| 68:20.08.08 Short-acting Insulins | Insulin Human |
| 68:20.08.16 Long-acting Insulins | Insulin Degludec |
|  | Insulin Degludec and Liraglutide |
|  | Insulin Detemir |
|  | Insulin Glargine and Lixisenatide |
|  | Insulin Glargine |

### Methods Supplement

This methods supplement, which accurately describes the data used in this manuscript, was duplicated from the following paper using the same data:

Zimmerman SC, Ferguson EL, Choudhary V, et al. Metformin Cessation and Dementia Incidence. JAMA Netw Open. 2023;6(10):e2339723. doi:10.1001/jamanetworkopen.2023.39723

### Diabetes registry

Individuals meeting any one of four criteria were considered to have diabetes and included in the registry: (1) a single, principal inpatient diabetes mellitus (DM) diagnosis; (2) two or more outpatient diabetes diagnoses (ICD-9 and ICD-10 codes) during the previous 5 years; (3) two or more abnormal outpatient lab test results during the previous two years (i.e., HbA1c ≥ 6.5\%, fasting glucose ≥ 126 mg/dL, random glucose or post-load glucose ≥ 200 mg/dL); or (4) at least one dispensing for a medication used to treat DM (insulin, oral hypoglycemic agent, antihyperglycemic agents). Individuals who use metformin or thiazolidinediones and have certain diagnoses (i.e., polycystic ovary syndrome) during the past 2 years, but did not meet any of the other diabetes criteria were excluded from the registry. Patients with evidence of gestational diabetes are also excluded from the registry.

### Renal registry

Ages of dialysis and kidney transplant events were data were extracted from the KPNC renal registry.

### Prescription Records

Prescriptions of metformin, insulins, and other antidiabetes drugs were extracted from KPNC pharmacy dispensing records by generic name. Periods of continuous medication use were calculated without respect to dosage. Each prescription record included a start date and a number of days’ supply. Prescription end dates were calculated by adding the days’ supply to the start date, and prescriptions were collapsed into contiguous periods if the gap between the end of a prescription and the start of the next prescription was 182 or fewer days (“persistence requirement”). For collapsed prescriptions, adherence was calculated as the number of days of medication supply divided by the remaining days on the prescription. A new prescription record was created if adherence fell below 0.8. For combination medications, separate records were included for each component drug. Details on prescriptions are given in appendix table 1.

### Diagnosis Records

We determined diabetes status (type 1, type 2, or neither) from the KPNC Diabetes Registry. The registry has records dating back to 1978 for any encounter type, allowing for ascertainment of diabetes diagnosis beyond what would be available in electronic health records. Additional information on the registry is available in the Supplement and citation below. We defined clinical diagnoses of cardiovascular disease, cancer, kidney disease, and dementia in terms of groups of International Classification of Diseases, Ninth Revision (ICD-9) diagnostic codes for January 1996 to September 2015 and International Classification of Diseases, Tenth Revision (ICD-10) diagnostic codes after September 2015. The outcome measure of all-cause dementia diagnosis was defined as first diagnosis of any of the following: Alzheimer’s disease, vascular dementia, non-specific dementia, dementia with Lewy bodies, and Parkinson’s disease. All diagnostic codes are given in appendix table 1.

Moffet, H.H., Adler, N., Schillinger, D., Ahmed, A.T., Laraia, B., Selby, J.V., Neugebauer, R., Liu, J.Y., Parker, M.M., Warton, M. and Moffet, H.H., 2007. The Diabetes Study of Northern California (DISTANCE): Objectives and design of a survey follow-up study of social health disparities in a managed care population.

### Lab Records

Glycated hemoglobin, estimated glomerular filtration rate (eGFR), and serum creatinine test results were extracted from KPNC lab records.

### Sociodemographic Variables

Gender was extracted from KPNC health plan membership databases. Other sociodemographic covariates (race, ethnicity, educational attainment, nativity, parental nativity, and survey language) were self-reported in the CMHS or RPGEH surveys. Self-reported race and ethnicity categories were collapsed into the following categories: Black, Hispanic/Latino, Asian, White and other/uncertain. The CMHS and RPGEH surveys asked participants to endorse specific racial/ethnic identities (e.g., African-American, Mexican, Cuban, Chinese, Japanese, White or European-American, etc.). Participants were instructed to endorse all groups that applied. For individuals who endorsed more than one race or ethnicity, these indicators were collapsed using an algorithm to assign one adjudicated race/ethnicity with the following prioritization order: Black, Hispanic, Asian, Native American (with no other racial/ethnic group endorsement). If a participant did not endorse any ethnicities on their survey, race/ethnicity was derived from the KPNC health plan databases. The algorithm was validated by comparing against several sources, including the KPNC virtual data warehouse (VDW), Utility for Care Data Analysis (UCDA; a Kaiser division that collects and cleans demographic data on Kaiser membership), and principal components from genetic data (available for ∼25% of participants). The majority of the full RPGEH cohort had race/ethnicity values from the survey that matched these other sources and genetic data when available. For the small percentage (∼2%) that did not match, surveys were checked by hand (to account for possible scanning errors), and race/ethnicity endorsement(s) were overwritten with VDW data when errors were found.

### KPNC Membership

Start and end ages of each period of KPNC membership for each person were extracted from an administrative database. Subsequent KPNC membership periods were collapsed if the duration of a membership gap was 90 days or shorter.

### Death Records

Death dates were obtained from the KPNC mortality database, which combines KPNC clinical and administrative, National Death Index, California State death, and Social Security Administration records.

### Censoring

Each participant was assigned an “administrative censoring” date randomly chosen between January 30th, 2020 and March 31st, 2020 so that date of birth could not be inferred in the analytic dataset. To protect participant anonymity, participants in the dataset were topcoded at age 90, therefore all participants were censored upon reaching age 90.

